# Epigenome-wide characterization reveals aberrant DNA methylation of host genes regulating CD4+ T cell HIV-1 reservoir size in women with HIV

**DOI:** 10.1101/2024.07.26.24311074

**Authors:** Ke Xu, Xinyu Zhang, Kesava Asam, Bryan C. Quach, Grier P. Page, Deborah Konkle-Parker, Claudia Martinez, Cecile D. Lahiri, Elizabeth T. Topper, Mardge H. Cohen, Seble G. Kassaye, Jack DeHovitz, Mark H. Kuniholm, Nancie M. Archin, Amir Valizadeh, Phyllis C. Tien, Vincent C. Marconi, Dana B. Hancock, Eric Otto Johnson, Bradley E. Aouizerat

## Abstract

The underlying mechanism of the HIV-1 reservoir, a major barrier to an HIV cure, is largely unknown. The integration of HIV-1 DNA and immune defense mechanisms can disrupt the host epigenetic landscape, potentially silencing HIV-1 replication. Using bisulfite capture DNA methylation sequencing, we profiled approximately 3.2 million CpG sites in CD4^+^ T cells isolated from the blood of 427 virally suppressed women with HIV. The average total CD4^+^ T cell HIV-1 Reservoir (HR_CD4_) size was 1,409 copies per million cells. Most proviruses were defective with only a small proportion being intact. We found 245 differentially methylated positions (CpG sites) and 85 methylated regions associated with the total HR_CD4_ size. Notably, 52% of significant methylation sites were in intronic regions. HR_CD4_-associated genes were involved in viral replication (e.g., *ISG15*), HIV-1 latency (e.g., *MBD2*), and cell growth and apoptosis (e.g., *IRF9*). A subset of the identified genes with aberrant methylation was an established target of HIV-1 integration (e.g., *NFIA, SPPL3, DLEU2, ELMSAN1*). Overall, HR_CD4_ size was inversely associated with DNA methylation of interferon signaling genes and positively associated with methylation at established HIV-1 integration sites. HR_CD4_-associated genes were enriched in pathways including immune defense against the virus (i.e., interferon-α response and interferon-γ response), DNA binding transcription repression, and host-virus interaction such as Tau protein binding. Together, our results show that epigenomic alterations in CD4^+^ T cells are associated with total HIV-1 reservoir size, offering new insights into HIV-1 latency and may provide potential molecular targets for future HIV-1 eradication strategies.

## Introduction

The HIV-1 reservoir (HR) has posed a major barrier to an HIV cure. HIV latency is established early during infection, with the provirus persisting primarily in resting or memory CD4^+^ T cells^1,2^. The HR evades eradication by the immune system and is largely suppressed by antiretroviral therapy (ART). However, cessation of ART leads to HIV-1 rebound within approximately 2 weeks^3^. The HR size decays slowly with an estimated half-life of 44 months for the total HR^4^ and 59 months for the intact provirus^5^. At this decay rate, eradication would take over 60 years, necessitating lifelong ART for people with HIV (PWH).

In recent years, efforts to cure HIV have focused on latency reversal followed by immune-mediated clearance of infected cells. However, latency reversal agents have failed to effectively reduce HR size, presenting a major challenge^6,7^. Another challenge is the limited understanding of how HR establishes, maintains, and rebounds in CD4^+^ T cells before, during, and after ART discontinuation.

Emerging evidence shows that epigenetic regulation of both the viral and host genomes is crucial to establishing and maintaining HIV-1 latency. Transcription of provirus depends upon the dynamic interaction between genomic alteration of the provirus and host immune activity^8^. The HIV-1 provirus genome reduces chromatin accessibility in latently infected cells^9^. Mechanistically, two CpG sites in the promoter region of long terminal repeats (LTR) of the provirus are hypermethylated, mediated by the host methyl-CpG binding domain protein 2 (MBD2) in infected CD4^+^ T cells^10^. In contrast, some studies reported that methylation of LTR was not associated with HIV-1 latency^11,12^. Research on the host genome’s DNA methylation and HR is still in its infancy. Some studies have reported that hyper- and hypo-methylated regions are associated with HR size and positively correlate with disease progression assessed by viral response to ART^13^, but these findings are based on small sample sizes or *ex vivo* experiments, making it challenging to draw strong conclusions about *in vivo* host DNA methylation dynamics associated with HR.

Genome-wide DNA methylation analysis with high-density CpG profiling is a useful approach to comprehensively investigate the role of the host DNA methylome in HR size. Among the total reservoir in infected CD4^+^ T cells, only a small proportion contains intact provirus (∼1 per million cells) capable of full transcription activation. Most proviruses are defective because of apolipoprotein B mRNA editing enzyme catalytic polypeptide-like 3G (*APOBEC3C*)-mediated hypermutation^14^, large internal deletions, packaging signal (Ψ) deletions, major splice donor mutations, and inactivating point mutations^15^. Most defective proviruses accumulate swiftly during early HIV-1 infection^14^. While previous studies have focused on intact reservoirs, recent research indicates that defective provirus can be activated to produce HIV-1 transcripts when integrated at active transcription sites^15^, and it can be recognized by cytotoxic T cells^16^. Proviruses escape mutations and remain stable during ART^17^, leading to chronic immune activation and inflammation, which are associated with the progression of HIV-related comorbidities^18–20^. Thus, it is plausible that host epigenome alterations occur in response to both intact and defective reservoirs after HIV-1 integration. Characterization of the perturbed host epigenome by intact and defective proviruses may help us discover new targets to eradicate the reservoir.

In this study, we examined the association of DNA methylation features with HR size in CD4^+^ T cells (HR_CD4_) from a cohort of women with HIV (WWH) who were virally suppressed on ART (N=427). DNA samples were extracted from isolated CD4^+^ T cells. Using a methylation capture sequencing method, we profiled DNA methylation sites across the epigenome and found 245 methylation sites associated with total HR_CD4_. The genes annotated for significant methylated sites were involved in HIV-1 latency, inflammation, and cell differentiation and harbored HIV-1 integration sites. These findings provide new evidence on the potential roles of host epigenetic factors in HR size through type I interferon genes, immune activation, and chromosome stability.

## Results

### Participant characteristics and HIV-1 latent reservoir detection

The study population was derived from the Women’s Interagency HIV Study (WIHS) a multi-center interval cohort study of women with HIV (WWH) and a comparison group of women without HIV infection in the United States^21,22^. In 2019, WIHS merged with the Multicenter AIDS Cohort Study (MACS) to form the MACS/WIHS Combined Cohort Study (MWCCS)^23^. Participants underwent follow-up visits every six months which included the collection of peripheral blood mononuclear cells (PBMC) and demographic, clinical, and ART treatment data. The 427 participants who met the inclusion and exclusion criteria were selected for the current study (See **Methods**). The average age was 47.3 ± 8.2 years; 76% of the participants self-identified as African American non-Hispanic, 12% as White non-Hispanic, and 11% as other races. Most participants (80%) currently smoked tobacco. All participants were on ART and had undetectable viral loads (below the assay lower limit of quantitation) for a minimum of six months, with an average duration of 2.1 years, up to the visit at which CD4+ T cells were isolated from PBMCs and were processed to estimate the HR_CD4_. The genomic DNA extracted from CD4+ T cells was subjected to bisulfite capture sequencing for DNA methylation profiling.

The HR_CD4_ quantification method for this cohort was detailed previously by Aouizerat et al.^24^. Briefly, HR_CD4_ was detected using droplet digital polymerase chain reaction (ddPCR). Intact proviral HIV DNA was measured using a modified intact proviral DNA assay (IPDA). The average HR_CD4_ size was 1,409 ± 1,639 copies per million CD4^+^ T cells. As expected, defective HIV-1 proviruses were more common than intact HR_CD4_. The average intact HR_CD4_ was 279 ± 463 copies per million; defective HR_CD4_ was 531 ± 886 copies per million for APOBEC3G-mediated mutations (3’- defective HR_CD4_) and 600 ± 748 copies per million for large internal deletions (5’- defective HR_CD4_). Demographic and clinical characteristics are presented in **Table 1**.

**Table 1.** Demographic and clinical characteristics of participants.

| Variable |  | Participants (N=427) |
| --- | --- | --- |
| Age (Mean ± SD) | | 47.29 ± 8.22 |
| Sex (F/M) |  | 427/0 |
| Ancestry (%) | African American | 76.3 |
|  | European | 12.4 |
|  | Other | 11 |
| Smoking Status (%) | Current | 79.9 |
|  | Former | 8.9 |
|  | Never | 11 |
| Alcohol Drinks per Week (Mean ± SD) | | 9.3 ± 26.5 |
| On ART + Undetected VL (%) |  | 100 |
| HLR (Mean ± SD) | Total | 1,409 ± 1,639 |
| | Intact | 279 ± 463.4 |
| | 5'-depletion | 600 ± 747.7 |
| | 3'-depletion | 530 ± 885.7 |

ART: Antiretroviral Therapy; HR: HIV-reservoir; VL: Viral load

### Epigenome-wide methylation characterization of HR_CD4_

CD4^+^ T cells epigenome was profiled using bisulfite methylation capture sequencing, yielding data on methylation of 3.2 million CpG sites. Quality control (QC) was conducted according to our previous report^25^ (see **Methods**). Only CpG sites with a coverage greater than 10x depth were included. Approximately 2 million CpG sites were analyzed after QC. Ancestry and chronological age were included in generalized linear regression models.

We found 245 differentially methylated sites associated with total HR_CD4_ size (False Discovery Rate, FDR<0.05) (**Figure 1A, Supplementary Table**). Among them, 67% (n=165) were positively associated with HR_CD4_ size while 33% (n=80) were negatively associated with it (**Figure 1B**). A substantial proportion of these methylation sites (89%) were annotated within or near genes, with the rest (11%) in intergenic regions. Within gene regions, 52% of CpG sites were in introns, 20% in promoter regions, and 11% in exons (**Figure 1C**). Additionally, 26% of the associated sites mapped to CpG islands, with the rest in non-island regions (open sea 34%, shore 30%, and shelf 9%). Notably, although most of the observed associations were positive, 50% of the methylation sites in the promoter regions were negatively associated with total HRCD4.

**Figure 1.**
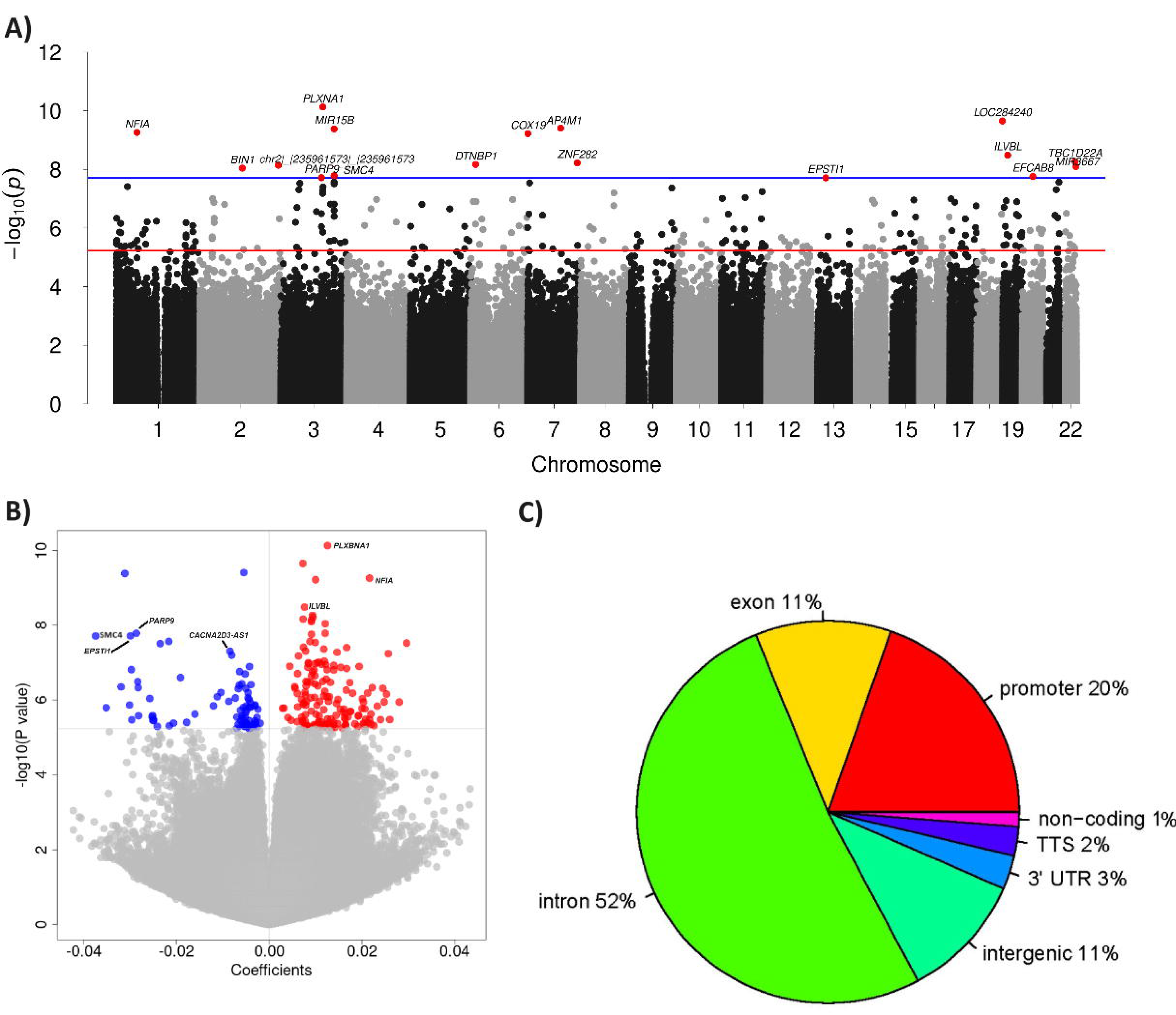
Epigenome-wide association analysis of CD4^+^ T cell total HIV-1 reservoir size: (A). Manhattan plot; (B) Volcano plot showing positive and negative association of CpG sites with total HR; (C) Proportion of CpG site annotations.

We further examined whether the total HR_CD4_-associated CpG sites were also significantly associated with intact and defective HR_CD4_. Among the 245 significant CpG sites for total HR_CD4_, 38 were nominally associated with intact or defective HR_CD4_ (p<0.05). For example, *SMC4* on chromosome 3 at position 160119770 was negatively associated with total (t=-5.79, p=1.64e-08), intact (t=-4.0, p=7.63e-05), and 5’-defective (t=-4.78, p=2.64e-06) HR_CD4_. *PARP9* on chromosome 3 at position 122281940 was also negatively associated with total (t=-5.76, p=1.92e-08), intact (t=-3.19, p=1.57e-03), 5’- defective (t=-3.66, p=2.93e-04), and 3’-defective (t=-4.28, p=2.39e-05) HR_CD4_. *ISG15* on chromosome 1 at position 949432 was negatively associated with total (t=-5.14, p=4.66e-07), intact (t=-3.39, p=7.90e-04), and 5’-defective (t=-4.53, p= 8.24e-06) HR_CD4_. These genes were previously reported to be involved in interaction with HIV-1 tau protein (*SMC4*)^26^, HIV-1 induced apoptosis (*PARP*)^27^, and HIV-1 replication (*ISG15*)^28^, which may contribute to HIV-1 latency.

### Differentially methylated regions were significantly associated with total HR_CD4_

To find significant differentially methylated regions (MR), we conducted an analysis using the “bump hunting” approach (see **Methods**). This analysis found MRs containing at least 2 CpG sites associated with total HR_CD4_. We found 85 significant MRs across 22 chromosomes (**Table 2**). Among them, 28 MRs were negatively associated with HR_CD4_ while 57 MRs were positively associated.

**Table 2.** Differentially methylated regions associated with total HIV-1 reservoir in CD4^+^ T cells.

| Chr | Start | End | Gene | # CpGs | t | p | FDR |
| --- | --- | --- | --- | --- | --- | --- | --- |
| 1 | 61545077 | 61545123 | NFIA | 2 | 5.4 | 1.1e-09 | 9.2e-08 |
| 3 | 160119633 | 160122505 | MIR16-2, MIR15B, SMC4 | 12 | -5.0 | 4.7e-09 | 2.0e-07 |
| 3 | 122281883 | 122281941 | DTX3L, PARP9 | 2 | -5.4 | 3.8e-08 | 1.1e-06 |
| 13 | 43565289 | 43565401 | EPSTI1 | 3 | -4.7 | 5.8e-08 | 1.2e-06 |
| 14 | 74250898 | 74250927 | ELMSAN1 | 2 | 4.8 | 2.9e-07 | 4.1e-06 |
| 3 | 54888035 | 54888057 | CACNA2D3 | 6 | -4.4 | 2.9e-07 | 4.1e-06 |
| 2 | 43398273 | 43398277 | - | 2 | 5.1 | 5.4e-07 | 6.5e-06 |
| 7 | 44924186 | 44924194 | PURB | 2 | -4.5 | 6.7e-07 | 7.1e-06 |
| 1 | 12676155 | 12676183 | DHRS3 | 2 | 5.0 | 9.4e-07 | 7.7e-06 |
| 19 | 41815093 | 41815101 | CCDC97 | 3 | 4.5 | 1.0e-06 | 7.7e-06 |
| 10 | 88852547 | 88852560 | GLUD1 | 2 | 4.6 | 1.1e-06 | 7.7e-06 |
| 1 | 119522623 | 119522631 | TBX15 | 2 | 4.5 | 1.1e-06 | 7.7e-06 |
| 1 | 949394 | 949433 | ISG15 | 3 | -4.6 | 1.2e-06 | 7.7e-06 |
| 11 | 118790174 | 118790223 | - | 3 | 4.6 | 1.4e-06 | 7.7e-06 |
| 1 | 207996299 | 207996461 | - | 3 | 4.8 | 1.4e-06 | 7.7e-06 |
| 17 | 7381527 | 7381577 | ZBTB4 | 2 | 4.5 | 2.4e-06 | 1.3e-05 |
| 21 | 42798945 | 42799143 | MX1 | 7 | -4.4 | 2.7e-06 | 1.3e-05 |
| 15 | 44829519 | 44829525 | EIF3J, EIF3J-AS1 | 2 | -4.5 | 3.0e-06 | 1.4e-05 |
| 1 | 207996636 | 207996676 | - | 2 | 4.7 | 3.3e-06 | 1.4e-05 |
| 9 | 134139856 | 134139880 | FAM78A | 3 | 4.2 | 3.3e-06 | 1.4e-05 |
| 10 | 22612799 | 22613027 | COMMD3-BMI1, BMI1 | 2 | 4.5 | 3.4e-06 | 1.4e-05 |
| 22 | 18635396 | 18635462 | USP18 | 3 | -4.4 | 3.8e-06 | 1.4e-05 |
| 16 | 4714768 | 4714796 | MGRN1 | 3 | 4.5 | 4.1e-06 | 1.4e-05 |
| 2 | 37551058 | 37551083 | - | 5 | 4.6 | 4.1e-06 | 1.4e-05 |
| 11 | 75062095 | 75062115 | ARRB1 | 2 | 4.4 | 6.3e-06 | 2.1e-05 |
| 21 | 42797710 | 42797717 | MX1 | 2 | -4.5 | 7.6e-06 | 2.4e-05 |
| 14 | 50159543 | 50159550 | KLHDC1 | 2 | 4.4 | 8.7e-06 | 2.6e-05 |
| 11 | 121324233 | 121324511 | SORL1 | 3 | 4.3 | 8.7e-06 | 2.6e-05 |
| 19 | 54876395 | 54876448 | LAIR1 | 2 | 4.2 | 9.3e-06 | 2.6e-05 |
| 1 | 12538489 | 12538543 | VPS13D | 4 | 4.4 | 9.3e-06 | 2.6e-05 |
| 1 | 949698 | 949878 | ISG15 | 12 | -4.2 | 1.1e-05 | 2.8e-05 |
| 1 | 949563 | 949599 | ISG15 | 4 | -4.3 | 1.1e-05 | 2.8e-05 |
| 15 | 69707926 | 69707938 | KIF23 | 2 | -4.4 | 1.2e-05 | 3.1e-05 |
| 17 | 47437978 | 47438153 | ZNF652, LOC102724596 | 3 | 4.2 | 1.4e-05 | 3.4e-05 |
| 15 | 55562701 | 55562788 | RAB27A | 4 | -4.1 | 1.7e-05 | 4.0e-05 |
| 22 | 27068060 | 27068092 | MIAT, MIATNB | 4 | 4.2 | 1.8e-05 | 4.2e-05 |
| 4 | 38666920 | 38667432 | KLF3-AS1, KLF3 | 2 | 4.3 | 1.8e-05 | 4.2e-05 |
| 12 | 120700363 | 120700438 | PXN | 3 | 4.0 | 2.4e-05 | 5.2e-05 |
| 9 | 6716463 | 6716482 | - | 4 | -4.2 | 2.5e-05 | 5.2e-05 |
| 17 | 79265630 | 79265645 | SLC38A10 | 2 | 4.1 | 2.5e-05 | 5.2e-05 |
| 4 | 103682822 | 103682824 | MANBA | 2 | 4.2 | 2.5e-05 | 5.2e-05 |
| 21 | 45509115 | 45509120 | TRAPPC10 | 2 | 4.3 | 2.7e-05 | 5.3e-05 |
| 11 | 614844 | 614857 | *IRF7* | 3 | -4.1 | 2.8e-05 | 5.4e-05 |
| 1 | 949479 | 949508 | *ISG15* | 3 | -4.2 | 2.8e-05 | 5.4e-05 |
| 10 | 90641813 | 90642005 | *STAMBPL1* | 3 | -4.2 | 2.9e-05 | 5.5e-05 |
| 14 | 105751631 | 105751638 | *BRF1* | 2 | 4.1 | 3.1e-05 | 5.6e-05 |
| 22 | 39637150 | 39637172 | *PDGFB* | 4 | 4.1 | 3.2e-05 | 5.6e-05 |
| 1 | 226069169 | 226069191 | *TMEM63A* | 2 | 4.1 | 3.2e-05 | 5.6e-05 |
| 19 | 4380572 | 4380611 | *SH3GL1* | 2 | 4.1 | 3.3e-05 | 5.6e-05 |
| 3 | 43385089 | 43385113 | *SNRK* | 2 | 4.2 | 3.4e-05 | 5.6e-05 |
| 14 | 24631374 | 24631383 | *IRF9* | 2 | -4.2 | 3.4e-05 | 5.6e-05 |
| 6 | 137630457 | 137630478 | - | 2 | 4.2 | 3.4e-05 | 5.6e-05 |
| 11 | 614567 | 614763 | *IRF7* | 2 | -4.2 | 3.7e-05 | 5.9e-05 |
| 9 | 6716396 | 6716425 | - | 5 | -4.1 | 3.9e-05 | 6.0e-05 |
| 17 | 42462262 | 42462268 | *ITGA2B* | 2 | 4.1 | 4.4e-05 | 6.7e-05 |
| 20 | 62530409 | 62530421 | *DNAJC5* | 2 | 4.1 | 4.7e-05 | 6.8e-05 |
| 2 | 43398215 | 43398245 | - | 2 | 4.1 | 4.7e-05 | 6.8e-05 |
| 2 | 173294414 | 173294494 | *ITGA6* | 2 | 4.0 | 4.8e-05 | 6.8e-05 |
| 21 | 45509391 | 45509401 | *TRAPPC10* | 3 | 4.1 | 4.8e-05 | 6.8e-05 |
| 22 | 38610341 | 38610350 | *MAFF* | 2 | 4.1 | 4.9e-05 | 6.8e-05 |
| 9 | 6716209 | 6716228 | - | 2 | -4.1 | 4.9e-05 | 6.8e-05 |
| 11 | 61213606 | 61213621 | *SDHAF2* | 2 | 4.1 | 5.2e-05 | 7.0e-05 |
| 11 | 74975246 | 74975248 | *ARRB1* | 2 | 4.0 | 5.3e-05 | 7.1e-05 |
| 17 | 7381831 | 7381857 | *ZBTB4* | 2 | 4.1 | 5.6e-05 | 7.3e-05 |
| 2 | 71681539 | 71681542 | *DYSF* | 2 | 4.0 | 6.1e-05 | 7.9e-05 |
| 19 | 19281257 | 19281272 | *MEF2BNB-MEF2B, MEF2B* | 2 | 4.0 | 6.3e-05 | 8.0e-05 |
| 11 | 109962729 | 109962762 | *ZC3H12C* | 2 | 4.0 | 6.8e-05 | 8.5e-05 |
| 21 | 45509454 | 45509457 | *TRAPPC10* | 2 | 4.0 | 6.8e-05 | 8.5e-05 |
| 14 | 61109826 | 61109857 | - | 2 | 4.0 | 7.1e-05 | 8.7e-05 |
| 4 | 85416937 | 85416953 | *NKX6-1* | 2 | 3.9 | 7.9e-05 | 9.5e-05 |
| 16 | 53535169 | 53535242 | *AKTIP* | 2 | 3.9 | 9.0e-05 | 1.1e-04 |
| 1 | 154318280 | 154318316 | *ATP8B2* | 2 | 3.9 | 9.7e-05 | 1.1e-04 |
| 8 | 133098472 | 133098504 | *HHLA1* | 2 | -3.9 | 1.1e-04 | 1.2e-04 |
| 17 | 37308830 | 37308847 | *PLXDC1* | 2 | 3.9 | 1.1e-04 | 1.3e-04 |
| 9 | 6716240 | 6716271 | - | 2 | -3.9 | 1.2e-04 | 1.3e-04 |
| 6 | 168714672 | 168714675 | *DACT2* | 2 | 3.9 | 1.3e-04 | 1.4e-04 |
| 9 | 6716324 | 6716351 | - | 3 | -3.9 | 1.3e-04 | 1.4e-04 |
| 9 | 79791171 | 79791179 | *VPS13A* | 2 | 3.9 | 1.4e-04 | 1.5e-04 |
| 3 | 122401131 | 122401302 | *PARP14* | 3 | -3.8 | 1.4e-04 | 1.5e-04 |
| 5 | 142435222 | 142435443 | *ARHGAP26* | 2 | -3.8 | 1.5e-04 | 1.5e-04 |
| 15 | 85177989 | 85178063 | *SCAND2P* | 2 | 3.8 | 1.5e-04 | 1.5e-04 |
| 1 | 79090602 | 79092806 | *IFI44L* | 2 | -3.8 | 1.5e-04 | 1.5e-04 |
| 21 | 38065618 | 38065639 | - | 2 | 3.8 | 1.9e-04 | 1.9e-04 |
| 5 | 17219328 | 17219331 | *BASP1* | 2 | 3.8 | 1.9e-04 | 1.9e-04 |

Notably, 4 MRs on *ISG15,* spanning 21 CpG sites, were significant, including one region harboring 12 CpG sites. All 4 regions on *ISG15* were inversely associated with HR_CD4_ (**Figure 2A**). These CpG sites were either located in the TSS region or exons, suggesting that the aberrant methylation of *ISG15* may regulate *ISG15* expression, which is associated with HR. Another large significant MR was located on chromosome 3 at *SMC4*, spanning 2,872 base pairs and harboring 12 CpG sites (**Figure 2B**).

**Figure 2.**
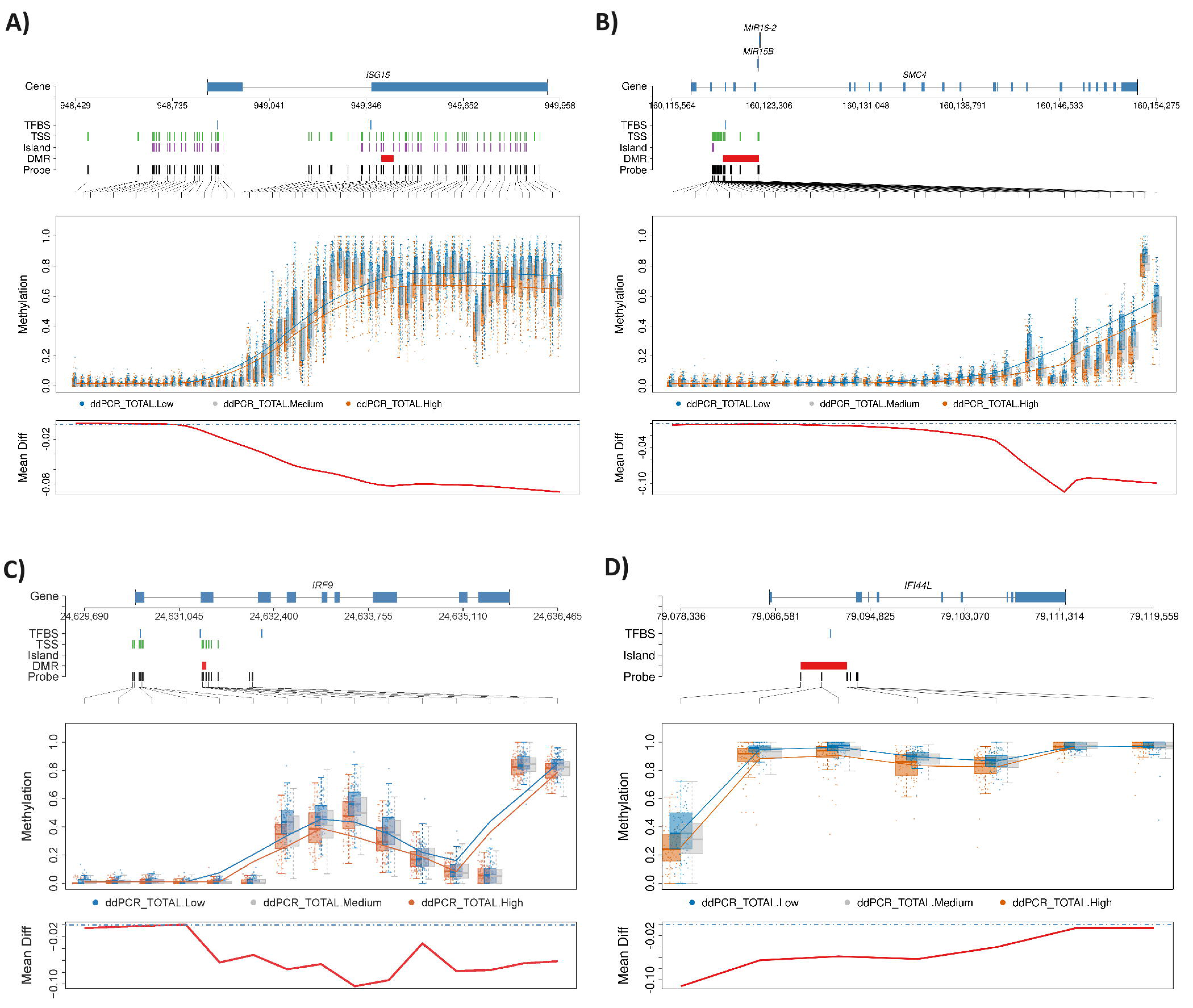
Differential methylation regions on four genes associated with HIV-1 reservoir size. (A) Interferon-stimulated gene 15 (*ISG15*); (B) Structural maintenance of chromosomes 4 (*SMC4*); (C) Interferon regulatory factor 9 (*IRF9*); (D) Interferon-induced protein 44 like (*IFI44L*). ddPCR- Total-low: bottom 25^th^ percentile HIV-1 reservoir size; ddPCR-Total-medium: medium HIV-1 reservoir size; ddPCR-Total-high: top 25^th^ percentile HIV-1 reservoir size.

Similarly, this MR was also inversely associated with HR_CD4_. Other negatively HR_CD4_- associated MR were in genes such as *PARP9, MX1, EPSTI1, USP18, IRF9, IFI44L,* and *IRF7* (**Figure 2C-D**).

### Total HIV-1 CD4+ T cell reservoir-associated DNA methylation is related to HIV-1 integration

Because half of the HR_CD4_-associated differentially methylated sites were located in intronic regions, we explored whether our 245 methylation sites were enriched in known HIV-1 genic integration sites by compiling HIV-1 integration sites reported across three recent studies^29–31^. We found that a subset of the genes harboring significant CpG sites were targets of HIV-1 integration. Methylation at most of these integration sites was positively associated with HR_CD4_ size.

One gene, *IMPDH2*, was a common HIV-1 integration site between our study and the three previous studies^29–31^. A CpG site on chromosome 3 at position 49066890, which was in the promoter region of *IMPDH2*, was positively associated with HR_CD4_ size. We also found that 23 HR_CD4_-associated genes overlapped with HIV-1 integration sites reported by Kok et al.^29^. Also, a few other genes were shared between our study and the other two previous studies^29,30^, including *NFIA, TMTC3, SPPL3, DLEU2, ELMSAN1, and ACSF3*. A total of 53 genes overlapped with HIV-1 integration sites reported by Maldarelli et al^31^ (e.g., *ISG15, TTC19, KCNG1, EIF3J, COX7A2L, TTC19, LAIR1, PARP9, JAK1,* and *CCDC97*), and 17 with sites reported by Wagner et al^30^ (e.g., *TTBK2, CDK16, ATP5PO,* and *TFDP1*) (**Figure 3**).

**Figure 3.**
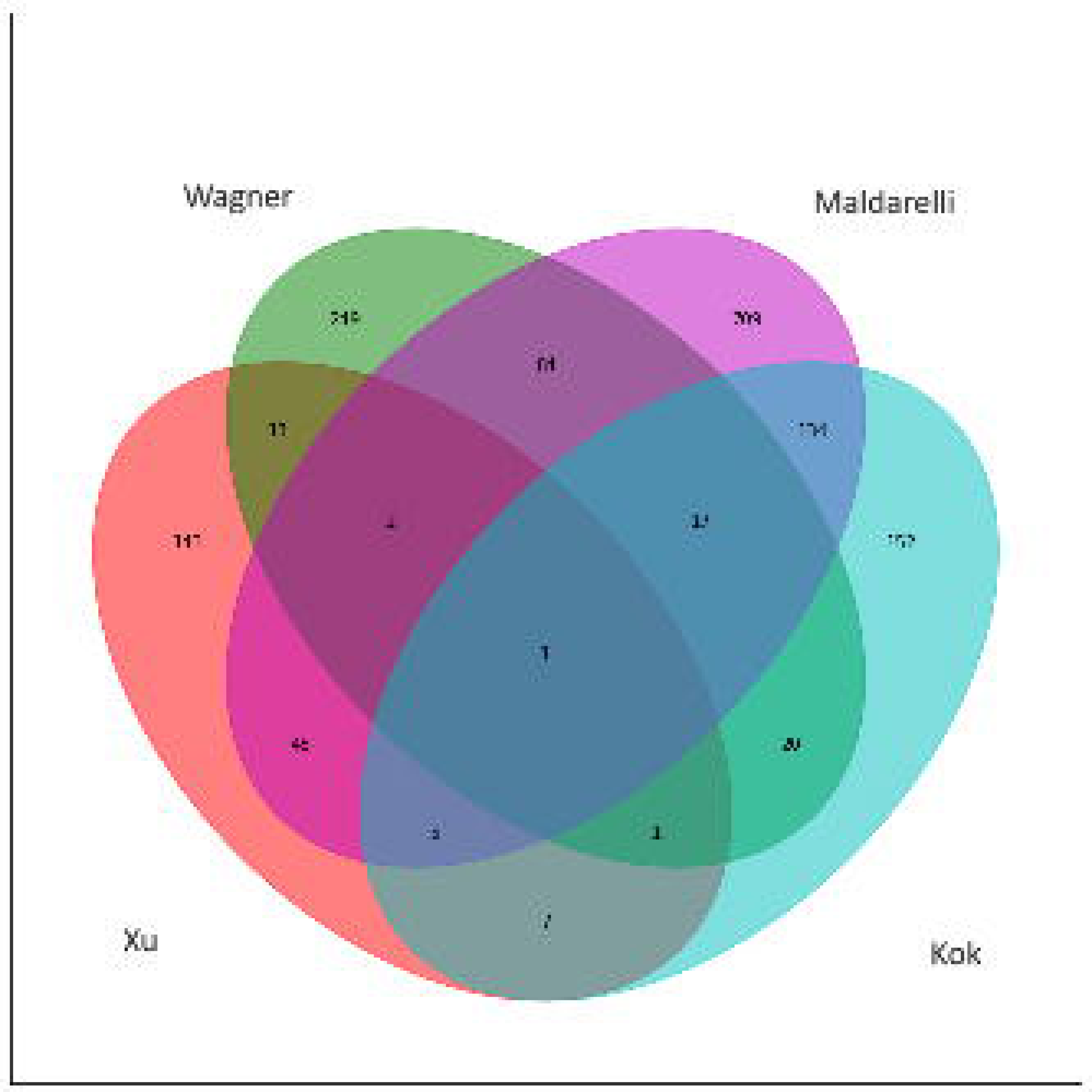
The number of significant genes associated with the HIV-1 reservoir found in this study (Xu) and its overlap with the previously identified HIV-1 integration sites in three other studies by Wagner et al, Maldarelli et al, and Kok et al.

The most frequently reported HIV-1 integration site, *BACH2*^30,31^, was nominally associated with HR_CD4_ (p=0.001) in our study. Additionally, several other previously reported integration sites displayed consistent patterns of differential methylation, with higher methylation levels in the top 25^th^ percentile of total HR_CD4_ size compared to the lowest 25^th^ percentile. For example, a CpG site at *JAK1* had 15% higher methylation in the top 25^th^ percentile for HR_CD4_ size, compared to the lowest 25^th^ percentile (*chr1 65503557,* p*=5.74e-07*) (**Figure 4A**). Similarly, two CpG sites at *NFIA* showed 7% and 10% higher methylation levels in the top 25^th^ percentile (*chr1 61542330,* p*=0.00004; chr1 61545122*, p*=5.49e-10)* (**Figure 4B**). Other examples include 3 CpG sites for *ELMSAN1 (chr14 74244367,* p*=6.62e-05; chr14 74250897,* p*=1.43e-07; chr14 74250926,* p*=0.00004) (***Figure 4C**), 2 CpG sites for *DLEU2 (chr13 50653658,* p*=0.00001; chr13 506554306,* p*=1.88e-06) (***Figure 4D**), and 1 CpG site for *SMAD2 (chr18 45458622:* p*=1.28e-05*) (**Figure 4E**).

**Figure 4.**
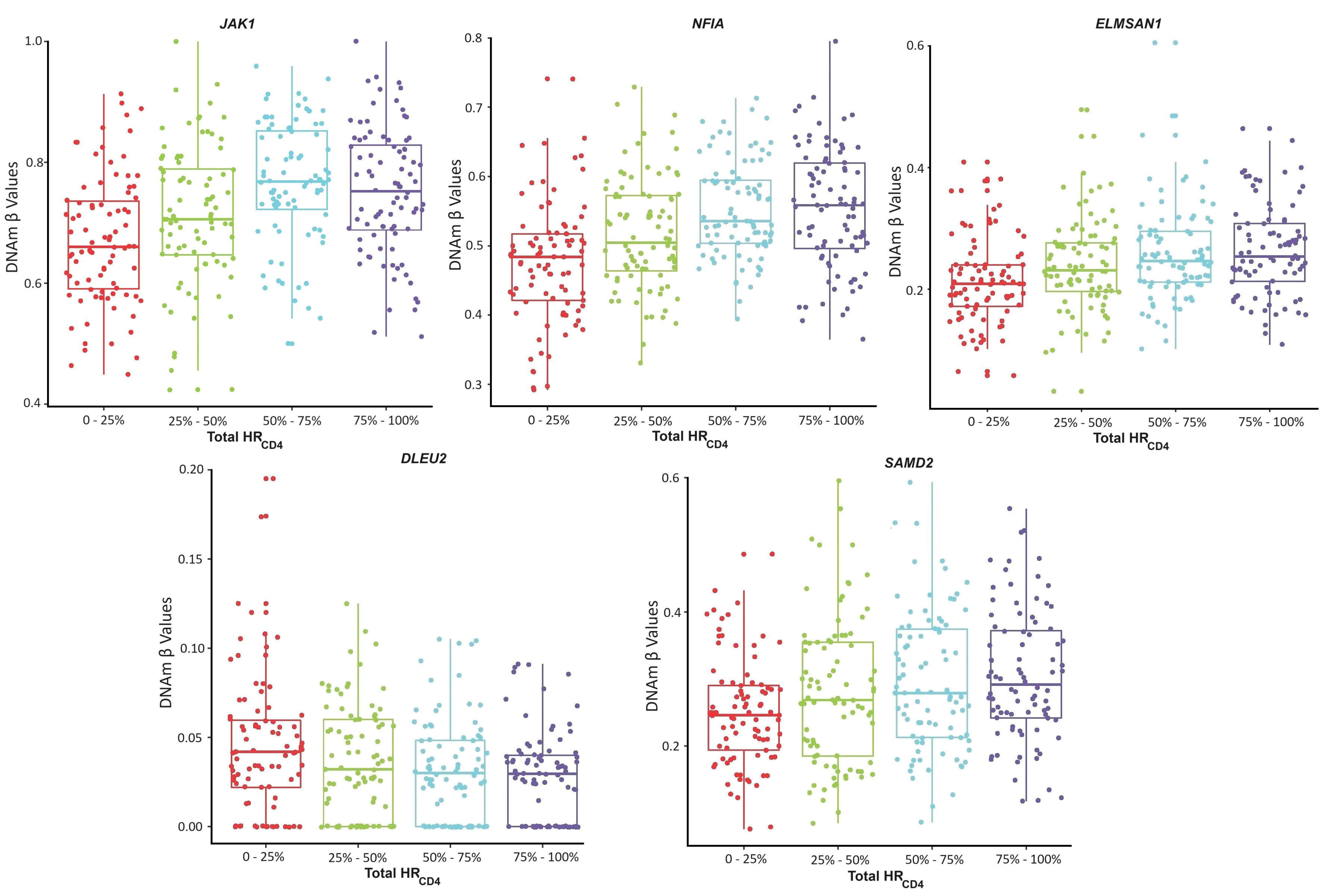
CpG sites differentially methylated between the high and low HIV-1 reservoir sizes on genes involved in HIV-1 integration sites. (A) *JAK1_chr1_6550357 p=5.74e-07; (B) two CpG sites on NFIA (chr1_6145122, p=5.49e-10 and chr1_61542330, p=0.00004); (C) three CpG sites on ELMSAN1(chr14 _74244367, p=6.62e-05; chr14 _74250897, p=1.43e-07; chr14_74250926: p=0.00004), and (D) two CpG sites on DLEU2 (chr13_50653658: p=0.00001 and chr13_506554306: p=1.88e-06); (E) SMAD2 chr18_45458622: p=0.0000128*

### Total HR_CD4_-associated CpG sites are enriched in pathways relevant to HIV-1 latency

We performed gene enrichment analysis using the human MSigDB database (https://www.gsea-msigdb.org/gsea/msigdb/), which includes 33,591 genes from nine major collections. In the hallmark gene set, we found significant enrichment for HR_CD4_- associated genes in the interferon α response (gene ratio: 16/79; p_adj_=1.50e-10) and interferon γ response (gene ratio: 16/79; p_adj_=3.45e-06) (**Figure 5A-B**) (**Table 3**). In the interferon α gene set, 10 genes were associated with HR_CD4_, with 8 of these overlapping between the “interferon α response” and “interferon γ response” gene sets. These shared genes were *USP18, MX1, ISG15, IRF7, IFIT3, IFI44L, IFI27*, and *EPST1*, all of which reached epigenome-wide significant association with total HR_CD4_. Four of these genes are involved in the regulation of inflammatory function (i.e., *IRF7, IFIT3, IFI44L, IFI27*).

**Figure 5.**
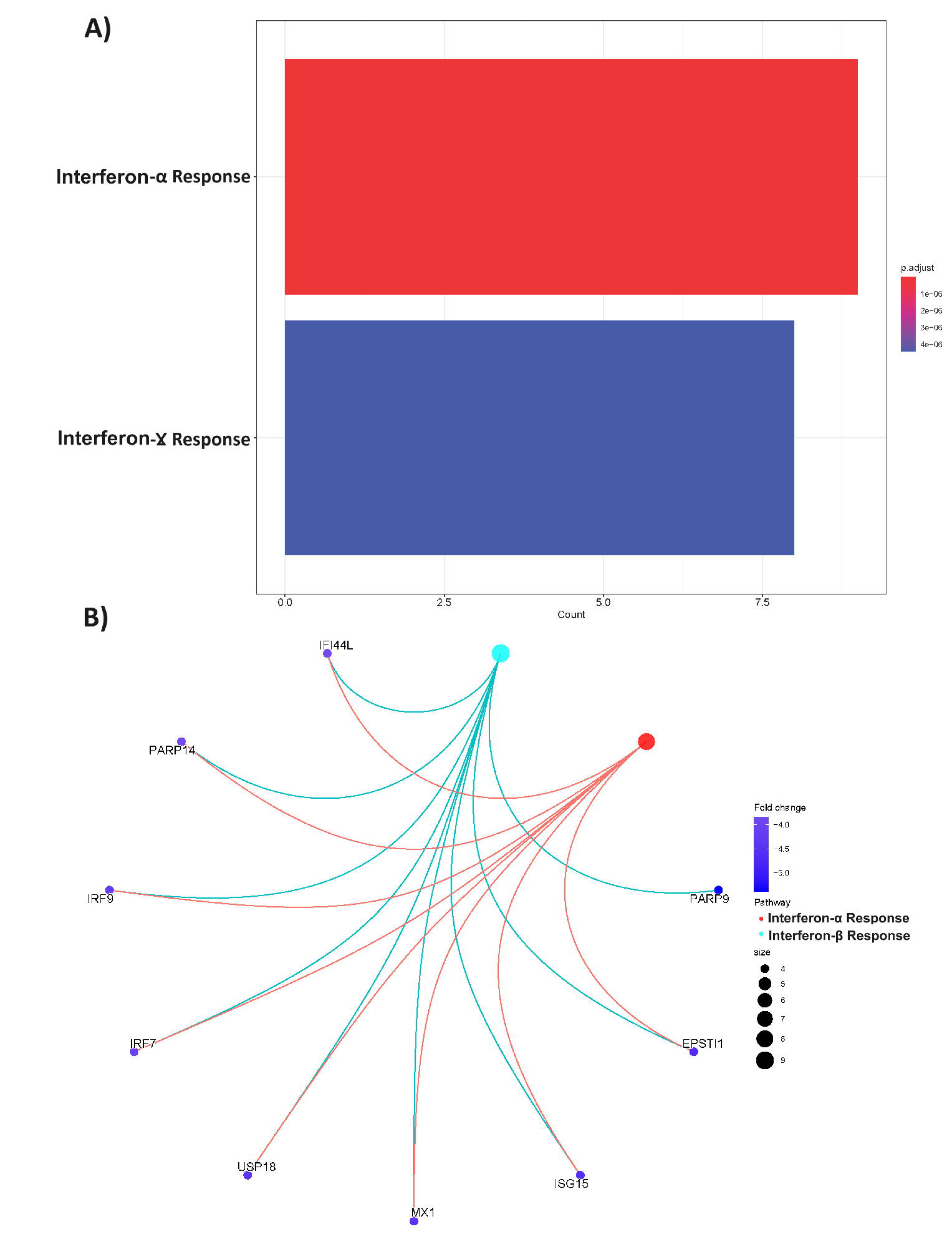
Significant pathways enriched on Hallmark gene set. (A) IFN-α and IFN-γ pathways; (B) HIV-1 reservoir-associated genes on those pathways.

**Table 3.** Significantly perturbed pathways associated with CD4+ T-cell total HIV-1 reservoir.

| Pathway | Gene Ratio | p | q |
| --- | --- | --- | --- |
| **Hallmark Pathways** |  |  |  |
| Interferon-α Response | 10/97 | 2.2e-05 | 1.0e-03 |
| Interferon-γ Response | 8/200 | 4.8e-02 | 7.7e-01 |
| **GO Molecular Function Pathways** |  |  |  |
| Tau Protein Binding | 5/272 | 1.1e-03 | 3.0e-01 |
| Vinculin Binding | 3/272 | 1.2e-03 | 3.0e-01 |
| DNA Binding Transcription Repressor Activity | 13/272 | 6.1e-03 | 6.7e-01 |
| DNA binding Transcription Activator Activity | 16/272 | 1.1e-02 | 6.7e-01 |
| DNA Binding Transcription Factor Binding | 16/272 | 1.2e-02 | 6.7e-01 |
| Aromatic Amino Acid Transmembrane Transporter Activity | 2/272 | 1.7e-02 | 6.7e-01 |
| Oligopeptide Transmembrane Transporter Activity | 2/272 | 1.7e-02 | 6.7e-01 |
| Intermediate Filament Binding | 2/272 | 2.0e-02 | 6.7e-01 |
| Peptide Transmembrane Transporter Activity | 2/272 | 2.0e-02 | 6.7e-01 |
| Amino Acid Transmembrane Transporter Activity | 5/272 | 2.0e-02 | 6.7e-01 |
| Transcription Factor Binding | 18/272 | 2.2e-02 | 6.7e-01 |
| Cell Adhesion Molecule Binding | 17/272 | 2.2e-02 | 6.7e-01 |
| Organic Anion Transmembrane Transporter Activity | 8/272 | 2.3e-02 | 6.7e-01 |
| 2-Oxoglutarate Dependent Dioxygenase Activity | 4/272 | 2.3e-02 | 6.7e-01 |
| Glycosyltransferase Activity | 10/272 | 2.3e-02 | 6.7e-01 |
| B-1-3-Galactosyltransferase Activity | 2/272 | 2.6e-02 | 6.7e-01 |
| Proton Transmembrane Transporter Activity | 6/272 | 2.7e-02 | 6.7e-01 |
| Glycosaminoglycan Binding | 9/272 | 2.9e-02 | 6.7e-01 |
| Mechanosensitive Ion Channel Activity | 2/272 | 3.0e-02 | 6.7e-01 |
| L-Amino Acid Transmembrane Transporter Activity | 4/272 | 3.2e-02 | 6.7e-01 |
| Demethylase Activity | 3/272 | 3.2e-02 | 6.7e-01 |
| Organic Acid Transmembrane Transporter Activity | 7/272 | 3.4e-02 | 6.7e-01 |
| Cadherin Binding | 11/272 | 3.4e-02 | 6.7e-01 |
| Insulin-Like Growth Factor Receptor Binding | 2/272 | 3.4e-02 | 6.7e-01 |
| Heparin Binding | 7/272 | 3.6e-02 | 6.7e-01 |
| Core Promoter Sequence-Specific DNA Binding | 3/272 | 3.7e-02 | 6.7e-01 |
| G-Protein Activity | 3/272 | 3.7e-02 | 6.7e-01 |
| Monosaccharide Binding | 4/272 | 3.8e-02 | 6.7e-01 |
| Magnesium Ion Transmembrane Transporter Activity | 2/272 | 3.8e-02 | 6.7e-01 |
| Protein Self-Association | 4/272 | 4.2e-02 | 6.7e-01 |
| NAD+ Binding | 2/272 | 4.2e-02 | 6.7e-01 |
| Opsonin Binding | 2/272 | 4.2e-02 | 6.7e-01 |
| Mannose Binding | 2/272 | 4.7e-02 | 6.9e-01 |
| Protein Serine Threonine Tyrosine Kinase Activity | 3/272 | 4.9e-02 | 6.9e-01 |

Gene set enrichment analysis employing Gene Ontology (GO) molecular function terms resulted in 34 HR_CD4_-associated gene sets that were nominally significant but did not remain significant after multiple-testing correction. Nevertheless, these gene sets were highly relevant to HIV-1 pathogenesis (**Figure 6A, B**) (**Table 3**). The top-ranked gene sets included “tau protein binding” (p=0.001), “vinculin binding” (p=0.001), “transcription factor binding”, “DNA binding transcription repressor activity” (p=0.006), “DNA binding transcription factor binding” (p=0.01), and “interferon binding” (p=0.02). Nine genes were enriched in more than one gene set (i.e., *SMAD2, FOXK2, FOXP1, IRX3, PURB, SK1, ISL1, NFIA, TFDP1*).

**Figure 6.**
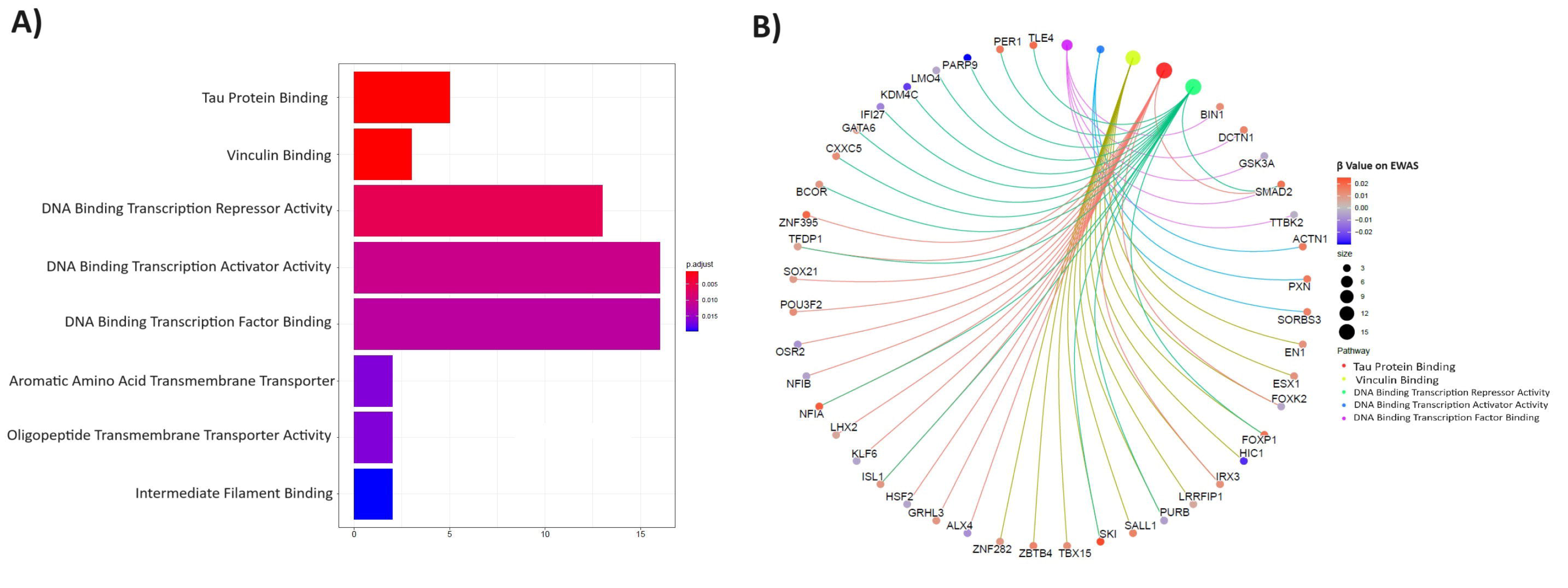
Gene set enrichment analysis results using GO Molecular function terms. (A) significant pathways; (B) HIV-1 reservoir-associated genes enriched on eight significant pathways; (C) the relationship between the top five pathways.

Interestingly, *SMAD2* connects the “tau-protein binding” pathway to the “DNA binding transcription factor” and “DNA binding transcription activator activity” pathways (**Figure 6C**). A methylation site at *SMAD2* was positively associated with HR_CD4_ (t=4.43, p=1.28e-05). *SMAD2* is responsible for transmitting the signal of transforming growth factor (TGF)-beta^32^, which plays a crucial role in regulating various cellular processes like cell proliferation, differentiation, and apoptosis.

## Discussion

Here, we report the first epigenome-wide association study on total HR_CD4_, highlighting significant new insights into DNA methylation mechanisms involved in the HIV-1 reservoir. Using bisulfite capture sequencing on samples collected from 427 women with virally suppressed HIV-1 infection, we comprehensively characterized the epigenomic landscape of HR_CD4_ over 2 million CpG sites. We found 245 DNA methylation sites and 87 regions associated with total HR_CD4_ size. Notably, genes and pathways related to significant CpG sites were involved in the host immune responses, particularly type I interferon genes and JAK1, which regulate HIV-1 replication and integration. We also observed HR_CD4_-associated aberrant methylation on the genes involved in chromosome stability, which may contribute to HIV-1 integration. Our results, derived from the largest sample size reported to date, reveal specific methylation mechanisms for HIV-1 persistence that may inform the potential target eradication strategies for HIV care.

Consistent with previous findings^15^, we found that the majority of the integrated DNA proviruses were defective. Recent studies have reported that both intact and defective integrated HIV-1 DNA proviruses remain transcriptionally active under suppressive ART, contributing to prolonged immune activation^33–35^ and greater comorbidity among PWH. Epigenetic regulation of host factors plays a critical role in HIV-1 transcriptome activation and silencing. In line with our current understanding of the mechanisms of HIV-1 pathogenesis, we found epigenetic alterations in genes involved in immune defense, interactions with viral proteins, and HIV-1 integration, suggesting that aberrant DNA methylation of these genes is linked to HIV-1 persistence. Notably, we found aberrant DNA methylation of multiple type I interferon genes associated with total HR_CD4_ size. For example, four significant CpG sites for the interferon-induced protein *ISG15* were inversely associated with HR_CD4_. A 445 base pairs region in *ISG15* containing multiple CpG sites was significantly less methylated in the highest total HR_CD4_ size quartile compared to the lowest quartile. Similarly, three regions in *interferon regulatory transcription factor 7 (IRF7)* encompassed 20 CpG sites that were less methylated in the highest compared to the lowest HR_CD4_ quartile. Additionally, a region spanning 4 CpG sites on *IRF9* was significantly less methylated in the highest compared to the lowest HR_CD4_ quartile. Other interferon-induced genes such as *IFI44L, MX1*, *PARP9,* and *USP18* consistently showed an inverse relationship between CpG methylation and HR_CD4_ size. Interferon genes are known to function as antiviral signaling molecules, regulate immune function, and induce apoptosis. As a result, it is presumed that increased expression of interferon genes due to hypomethylation may enhance restriction factor activity, resulting in limited HIV-1 replication and maintaining HR_CD4_ in a latent state. Furthermore, interferons also promote increased CD4+ T cell localization and HIV coreceptor expression, potentially enlarging the reservoir through seeding other cells. Our results support this complex and critical role of interferons in HIV-1 replication and latency^36^.

Another notable finding is that a hypermethylated CpG site at *JAK1* was associated with larger HR_CD4_ size. *JAK1* is a critical mediator of immune activation and is involved HIV-1 latency. Previous studies have shown that *JAK1* interacts with the HIV-1 transcriptome and is essential for its life cycle. HIV-1 envelop surface glycoprotein gp120^37,38^ has been found to induce the expression of *JAK1* through the Tat-mediated activity of PIK3R1^39^. Additionally, other studies have found that reduced expression levels of *JAK1* are associated with the inhibition of HIV LTR-beta-gal activity^40^. A clinical trial showed that Ruxolitinib, a JAK1/2 inhibitor, significantly decreased immune activation markers in PWH and reduced HIV-1 reservoirs^41^. Thus, the aberrant methylation of *JAK1* in relation to HR_CD4_ size in this study further supports the significant role of *JAK1* in HIV-1 latency.

HIV-1 genome tends to integrate into highly expressed genes and genes involved in clonal expansion and cell cycle control^29^. In this study, several genes involved in HIV-1 integration and latency showed significant association with HR_CD4_. For example, the methylation of *NFIA*, a transcription factor highly expressed in HIV-1 latently infected CD4+ T cells^42^, was positively associated with HR_CD4_ size, with the methylation being more pronounced in the highest HR_CD4_ size quartile.

Aberrant DNA methylation associated with HR_CD4_ was also discovered in genes responsible for chromosome stability, such as *SMC4*. *SMC4* is part of a family of genes that encode the structural maintenance of chromosome (SMC) proteins, which are parts of the cohesion and condensin complexes essential for chromatid pairing and chromosome segregation during mitosis^43^. Dysfunction of SMC proteins may increase vulnerability to HIV-1 integration into the host genome. They also play critical roles in regulating transcription and ATPase domains and are crucial for maintaining chromosome structure and the repair and stability of DNA. *SMC4* is also a positive regulator of innate inflammatory immune responses and cell proliferation^44^, which are key host factors for HIV-1 persistence. It is also believed that in T cells, *SMC4* interacts with HIV-1 Tat protein, which controls HIV-1 DNA replication and nuclear architecture^26^. The observed aberrant DNA methylation associated with HR_CD4_ size in *SMC4*, further supports its involvement in the establishment and maintenance of HR.

We acknowledge some limitations in our study. First, we were unable to find epigenome-wide significant methylation sites specific for intact or defective reservoirs, necessitating more sensitive detection methods. Additionally, sample input requirements and technical challenges of studying HR_CD4_ in a large sample of PWH precluded the measurement of the functional HR_CD4_ (e.g., QVOA) in conjunction with IPDA. A precise measurement of HIV-1 latency requires quantifying cellular HIV-1 RNA transcription. As a result, confirming the identified methylation sites for HR_CD4_ using single-cell methylation profiles is needed. In addition, we are unable to rule out the potential confounding effect of cigarette smoking on DNA methylation marks, as most of the participants were smokers. Lastly, our cross-sectional study design limits the ability to assess the causal relationship between altered DNA methylation and HR size. Both longitudinal and *ex vivo* studies are needed to figure out the causality between DNA methylation and reservoir size. Future research is also needed to replicate these findings in men living with HIV and explore the dynamic interaction between the genes found in this study and the HIV-1 genome in reservoir establishment and maintenance.

In summary, this comprehensive characterization of the epigenome concerning HR_CD4_ sheds light on the epigenetic mechanisms underlying HIV-1 persistence. Most importantly, these findings offer new gene targets such as *ISG15*, *PARP*, *NFIA,* and *SMC4* for an in-depth exploration of host responses to the HIV-1 reservoir. These targets could potentially serve as molecular points of interest for the development of novel treatments.

## Methods

### Ethics approval and consent to participate

The study was approved by the Committee of Human Research Subject Protection at Yale University and the Institutional Research Board Committee of the Connecticut Veteran Healthcare System. Informed consent was provided by all MWCCS participants via protocols approved by institutional review committees at each affiliated institution.

### Participants and HR detection

Clinical data and specimens used in this study were collected by MWCCS^23^. MWCCS is the largest observational cohort of HIV infection in the United States. The inclusion criteria were Women with HIV who had undetectable HIV-1 viral load (below the limit of quantitation for the assay) on ART treatment for at least 6 months^45^. A total of 427 WWH who met the criteria were selected for the study. The average duration of ART treatment was ∼2.1 years among the participants. CD4^+^ T cells were isolated by negative bead selection using the DynaBeads Untouched Human CD4 kit. Genomic DNA was then extracted from CD4^+^ T cells using magnetic bead-based nucleic acid isolation. ddPCR was employed to measure the intact proviral HIV DNA using the modified IPDA assay as described previously by Bruner *et al.*^46^, providing estimates of intact provirus per one millionCD4^+^ T cells. Additional details are discussed elsewhere^24^.

### Genome-wide DNA methylation capture sequence (MC-seq)

Genome-wide DNA methylation was conducted at the Yale Center for Genome Analysis according to the following steps:

### Methyl-Seq target enrichment library prep

Genomic DNA quality was determined by estimating the A260/A280 and A260/A230 ratios using spectrophotometry and the concentration from fluorometry. DNA integrity and fragment size were confirmed using a microfluidic chip run on an Agilent Bioanalyzer. Indexed paired-end whole genome sequencing libraries were prepared using the SureSelect XT Methyl-Seq kit (Agilent, part#G9651B). The library preparation was conducted following the manufacturer’s protocol. Briefly, samples yielding >350 ng were enriched for targeted methylation sites using the custom SureSelect Methyl-Seq Capture Library. The SureSelect enriched, bisulfite-converted libraries were PCR amplified using custom-made indexed primers (IDT, Coralville, Iowa). Dual-indexed libraries were quantified by quantitative polymerase chain reaction (qPCR) using the Library Quantification Kit (KAPA Biosystems, Part#KK4854). Insert size distribution was assessed using the Caliper LabChip GX system. Samples with a yield of ≥2 ng/ul proceeded to sequencing.

### Flow cell preparation and sequencing

Sample concentrations were normalized to 10 nM and loaded onto an Illumina NovaSeq flow cell at a concentration that yielded 40 million passing filter clusters per sample. Samples were sequenced using 100bp paired-end sequencing on an Illumina HiSeq NovaSeq, according to Illumina’s standard protocol. The 10bp dual index was read during additional sequencing reads that automatically followed the completion of the first read. Data generated during sequencing runs were transferred to the Yale Center for Genome Analysis high-performance computing cluster. A positive control (prepared bacteriophage Phi X library) provided by Illumina was spiked into every lane at a concentration of 0.3% to monitor sequencing quality in real-time.

### Preprocessing and quality control

Signal intensities were converted to individual base calls during a run using the system’s Real Time Analysis (RTA) software. Sample de- multiplexing and alignment to the human genome were performed using Illumina’s CASAVA 1.8.2 software suite. The sample error rate was required to be less than 1% and the distribution of reads per sample in a lane was required to be within reasonable tolerance. Quality control (QC) of sequencing reads was conducted following the standard procedure as previously described^47^. The quality of sequence data was examined using *FastQC* (ver. 0.11.8). Adapter sequences and fragments at 5’ and 3’ (phred score <30) with inadequate quality were removed by *Trim_galore* (ver. 0.6.3_dev). We used Bismark pipelines (ver. v0.22.1_dev) to align the reads to the bisulfite human genome (hg19) with default parameters^48^. An average of 87% of the sequence per sample was mapped to the genome. Quality-trimmed paired-end reads were transformed into a bisulfite-converted forward strand version (C->T conversion) or a bisulfite-treated reverse strand (G->A conversion of the forward strand). Only CpG sites with coverage >10 depth were used for final analysis to ensure the quality of CpG sites. A total of 427 samples and 2,078,054 CpG sites remained for epigenome-wide association analysis. Genes were annotated using Homer *annotatePeaks.pl*, including intergenic, 5’UTR, promoter, exon, intron, 3’UTR, transcription start site (TTS), and non-coding categories. CpG island, shore, shelf, and open sea annotation was defined by locally developed bash and R scripts based on genomic coordinates (hg19) of CpG islands from the UCSC genome browser. CpG shores were defined as up to 2 kb from CpG islands and CpG shelfs were defined as up to 2 kb from a CpG shore.

### Identification of CpG sites associated with HR

To ensure no other cell types were represented in the genomic DNA isolated from CD4^+^ T cells obtained from magnetic bead separation, we first estimated the PBMC cell type proportion using the Houseman method^49^. The distribution of the HIV-1 proviral DNA count (i.e., HR size) was normalized using a log10 transformation. Epigenome-wide association analysis was performed using generalized linear models, in which methylation beta value at each CpG site was modeled as the dependent variable, and total HR size was modeled as the independent variable, while also adjusting for age, self-reported race, cell type proportion, and smoking status. We also adjusted the model for the lower detection limit of each HIV-1 RNA viral load assay. The significance level was set at a false discovery rate (FDR) of 0.05. CpG sites that were evaluated for significant association with total HR were then re-evaluated for association with intact HR_CD4_, 5’-defective HR_CD4_, and 3’-defective HR_CD4_ separately.

### Identification of DNA methylation regions associated with total HR

Leveraging the bump-hunting framework^50^, we analyzed DNA methylation regions (as opposed to individual CpG sites) that were associated with total HR_CD4_. Bump hunting provides the advantage of effectively modeling measurement errors, removing batch effects, and detecting regions of interest for continuous measures such as HR_CD4_. We modified the bump hunting pipeline of Jaffe AE et al. to increase the sensitivity for detecting methylation regions. In the regression model, instead of using methylation β value at each CpG site, we used the *t*-statistic value that was derived from EWAS regressed on the total HR_CD4_. The p-value of each region was then estimated following the method proposed by Lui et al ^51^. A significant region was defined as having at least two consecutive CpG sites and an adjusted p<0.05.

### Gene set enrichment analysis

Genes adjacent to CpG sites were selected for gene set enrichment analysis. A cutoff of p<0.05 was used to ensure enough genes were selected for enrichment analysis. We used the Hallmark database from the Molecular Signatures Database (https://www.gsea-msigdb.org/gsea/msigdb/) and the Gene Ontology (GO) database for the enrichment analyses.

## Administrative information

### Author contributions

KX oversaw the study, including DNA methylation sequencing data collection, data processing, statistical analyses, and manuscript preparation. XZ conducted data processing and analysis and contributed to manuscript preparation. BEA and KA handled CD4^+^ T cell isolation, genomic DNA isolation, and HIV-1 reservoir detection. BCQ, GPP, and DBH participated in analytic strategy and manuscript preparation. DKP, HHB, CDL, ETG, MHC, NMA, MHK, and PCT participated in the study’s conduct, including protocol development, participant recruitment, follow-up, and biospecimen collection. AV contributed to manuscript preparation. VCM contributed to the interpretation of the findings and preparation of the manuscript. EOJ and BEA contributed to the study design, interpretation of findings, and manuscript preparation. All authors read and approved the final manuscript.

## Acknowledgments

The authors appreciate the support of the Yale Center of Genomic Analysis. Data for this study were collected by the Women’s Interagency HIV Study (WIHS), now the MACS/WIHS Combined Cohort Study (MWCCS). The authors gratefully acknowledge the contributions of the study participants and the dedication of the staff at the MWCCS sites.

## Data availability statement

Summary statistics of the epigenome-wide analysis are presented in the Supplementary Table. Access to individual-level DNA methylation data from the MACS/WIHS Combined Cohort Study Data (MWCCS) may be obtained upon review and approval of a MWCCS concept sheet. Links and instructions for online concept sheet submission are on the study website (https://statepi.jhsph.edu/mwccs/).

