## Supplementary figures and images for "Epigenome-wide characterization reveals aberrant DNA methylation of host genes regulating CD4+ T cell HIV-1 reservoir size in women with HIV"

### Supplementary Figure

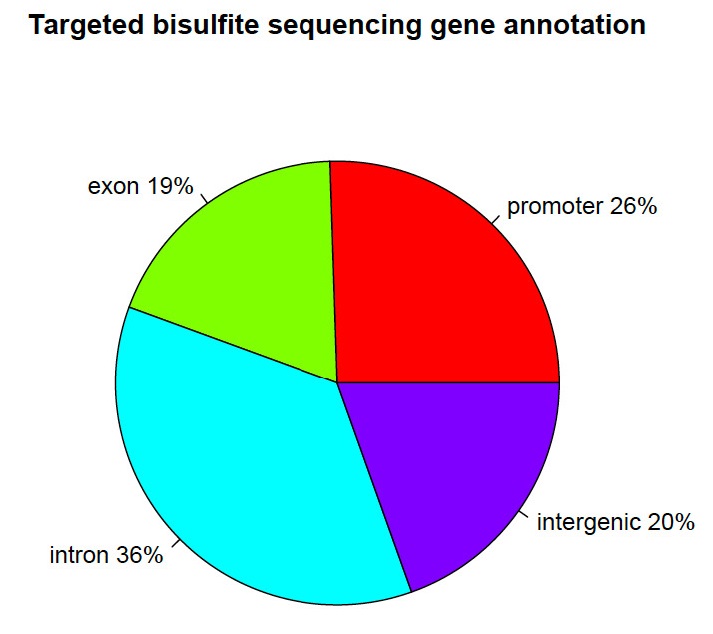
